# The myokine irisin represents an indirect pathway linking exercise to hippocampal subfields relevant to Alzheimer’s disease and neurogenesis

**DOI:** 10.1101/2025.09.09.25335471

**Authors:** Thomas Pace, Jacob M Levenstein, Bonnie L Quigley, Rhys Houston, Ana P Bouças, Sophie C Andrews

## Abstract

**INTRODUCTION:** While exercise is shown to reduce hippocampal atrophy, the underlying molecular mechanisms remain to be elucidated. Animal studies suggest the myokine irisin underlies exercise-related hippocampal benefits, though human evidence is lacking.

**METHODS:** We cross-sectionally examined 74 healthy older adults (age 65.47±8.56 years). Participants completed Godin Leisure-Time exercise questionnaires, provided fasting blood for irisin measurement, and underwent structural MRI with hippocampal subfield segmentation. Hierarchical regression and mediation analyses tested irisin-mediated exercise-hippocampus relationships, controlling for age, sex, and education.

**RESULTS:** Exercise positively associated with circulating irisin (β=0.365, p=0.003). Irisin positively associated with bilateral hippocampal volumes (right: β=0.353, p=0.001; left: β=0.275, p=0.012), strongest in right-CA3 (β=0.530), right-CA4/dentate gyrus (β=0.471), and bilateral CA1 (β=0.336-0.373) subfields. Mediation analysis revealed all exercise-hippocampus relationships operated indirectly through irisin.

**DISCUSSION:** This study provides first human evidence that irisin is a molecular mechanism linking exercise to hippocampal volume, particularly in subfields critical for memory, neurogenesis, and Alzheimer’s pathology.

## 1 BACKGROUND

Brain ageing is characterised by progressive structural and functional changes affecting cognition across the lifespan. In pathological ageing, these changes underpin dementia, with prevalence rates projected to triple by 2050 (1). Crucially, nearly half of dementia cases could be preventable through modifiable risk factors (2), with physical inactivity a key target. However, the molecular pathways through which physical activity confers neuroprotective effects on the human brain have not yet been fully elucidated.

Among brain regions, the hippocampus shows particular age-related vulnerability (3–7). Hippocampal atrophy begins in the sixth decade (4, 8) with ∼1.5% volume decline annually, accelerating to ∼5% pathologically (9–11), reaching ∼24% reduction in Alzheimer’s disease (AD) (12). Importantly, the hippocampus has a unique capacity for neurogenesis (13, 14) making it a promising therapeutic target (15), particularly via physical activity interventions (16).

Indeed, objective aerobic fitness scores correlate with larger hippocampal volumes cross-sectionally (17), while self-reported physical activity predicts preserved volumes over 9-12 years longitudinally (18, 19). Erickson et al. (2011) first provided causal evidence, demonstrating that a walking intervention reversed 1-2 years of age-related hippocampal loss in older adults (20). Meta-analyses have since reported reliable effects of exercise on hippocampal volume (21, 22), primarily through preventing age-related atrophy. However, the neurobiological mechanisms mediating exercise-induced hippocampal volume preservation remain unclear. One proposed mechanism involves myokine signalling.

During exercise, contracting skeletal muscle releases an array of signalling molecules called myokines into the bloodstream, which can cross the blood-brain-barrier and potentially influence neural tissue (23). Among exercise-induced myokines, irisin has become a popular candidate for explaining the positive effects of exercise on brain health (24–33), though definitive evidence in humans remains elusive. Discovered by Boström et al. (2012), irisin is a hormone-like myokine produced when exercise induces PGC-1α, leading to FNDC5 protein expression in skeletal muscle (34). FNDC5 is then proteolytically cleaved to release irisin into blood circulation.

Recently, animal studies have demonstrated irisin’s multiple roles in exercise-induced hippocampal health benefits. In rodent models, exercise increases hippocampal FNDC5/irisin expression by approximately 30-40% with region-specific effects (35–37). Direct administration of irisin or FNDC5 overexpression increased brain-derived neurotrophic factor (BDNF) expression, both *in vivo* and in cultured hippocampal neurons (35, 38). Further, irisin crosses the blood-brain-barrier (39), is necessary for hippocampal neurogenesis (40), and protects against neuronal apoptosis and inflammation (41). Additionally, genetic deletion of FNDC5/irisin impaired hippocampus-dependent memory and synaptic plasticity, while direct irisin administration rescued these deficits (39, 42). These animal studies have prompted human investigations.

Importantly, circulating irisin is elevated by acute exercise in humans as measured in serum and plasma (43–47), and is detectable in human cerebrospinal fluid (CSF) (48). In human neural cell culture models of AD, irisin promotes astrocytic secretion of the amyloid-beta (Aβ)-degrading enzyme neprilysin (49), and is reduced in the hippocampi and CSF of AD patients (42). While circulating irisin has been associated with better cognitive function in older adults and athletes (50–52), its relationship with human brain structure remains largely unexplored.

Only three neuroimaging studies have investigated irisin in humans. One found positive correlations between irisin and striatal dopamine uptake in Parkinson’s disease (53), while another found irisin associated with reduced cerebral small vessel disease burden in healthy men (54). Interestingly, the study that examined hippocampal volume found no association in healthy older adults (n=23) but negative associations in AD (51), potentially reflecting compensatory upregulation. To date, no study has demonstrated that higher irisin levels are associated with larger hippocampal volumes in healthy ageing, and critically, none have simultaneously measured exercise, irisin, and hippocampal volumes to test whether the compelling mechanistic pathway from animal models extends to humans. Therefore, this study provides the first investigation of exercise, irisin and hippocampal volumes in 74 healthy older adults. We hypothesised: (1) exercise levels are positively associated with circulating irisin; (2) exercise is associated with larger hippocampal volumes; (3) higher irisin relates to larger hippocampal volumes, and (4) irisin mediates the relationship between exercise and hippocampal volume.

## 2 METHODS

### 2.1 Participants & Procedure

This investigation utilised baseline data from participants enrolled in the Lifestyle Intervention Study for Dementia Risk Reduction (LEISURE) (55). Ethical approval for the LEISURE study was obtained from the Human Research Ethics Committee of the University of the Sunshine Coast (A191301). Written informed consent was secured from all participants prior to data collection. Eligible participants were healthy adults aged 50-84 years residing independently in the Sunshine Coast region of Queensland, Australia. Individuals were excluded if they had received diagnoses of mild cognitive impairment, dementia, traumatic brain injury, stroke, cardiovascular disease, metabolic disorders, serious psychiatric conditions, neurological disorders, epilepsy, diabetes, or stroke history.

From the 99 community-dwelling older adults who completed the initial LEISURE assessment protocol, 74 participants (females = 61, mean age = 65.47, SD ±8.56) had complete datasets across all variables of interest: blood biomarkers, neuroimaging, and exercise questionnaires.

The baseline assessment protocol comprised comprehensive medical, neuropsychological, and neurobiological evaluations conducted over multiple sessions at the Thompson Institute, University of the Sunshine Coast.

For the current investigation, relevant assessments included fasting blood sample collection for irisin analysis, structural MRI brain imaging, and completion of self-report questionnaires. All procedures were conducted by trained research personnel following standardised protocols. Participants completed assessments prior to randomisation and remained blinded to treatment allocation during baseline data collection. The full LEISURE protocol has been described in detail elsewhere (55).

### 2.2 Measures

#### 2.2.1 Circulating irisin levels

Irisin was measured from fasting serum samples. Whole blood was collected by a certified phlebotomist and serum was separated by centrifugation for 15 minutes at 2500 x g and 4°C. Samples were processed within 4 hours of collection and aliquots were stored at -80°C. Serum irisin levels were measured in duplicate and averaged using the Irisin ELISA kit (Human, Rat, Mouse, Canine; Phoenix Pharmaceuticals, California, USA) following the manufacturer’s instructions. All study samples yielded detectable levels of irisin within the kit’s quantifiable range (1044 ng/mL to 0.066 ng/mL).

#### 2.2.2 MRI Acquisition and Hippocampal volumes

All MRI brain scans were performed at the Nola Thompson Centre for Advanced Imaging (Thompson Institute, UniSC) using a 3T Siemens Skyra (Erlangen, Germany) and a 64-channel head and neck coil. With respect to the analysis performed, the following two anatomical scans were acquired: i. T1-weighted magnetization-prepared rapid gradient echo (MPRAGE: TR =2200ms, TE=1.71ms, TI=850ms, flip angle=7°, voxel resolution=1mm^3^, FOV=208×256×256, PAT-GRAPPA=2, TA=3:57), and ii. T2-weighted (TR=3200ms, TE=412ms, voxel resolution=1mm^3^, FOV= 240×256×256, PAT-GRAPPA=6, TA=2:54).

Prior to image processing, all anatomical brain scans were visually inspected for image quality, assessing image contrast, field homogeneity, head motion, image artefacts and field of view. No scans were removed due to poor image quality. Individuals’ T1-weighted scans were processed using FastSurfer’s FastSurferCNN (version 2.4.2; (56, 57)). As the primary analysis pertains to the hippocampus, visual inspection of FastSurferCNN’s segmentations were conducted on the aparc.DKTatlas+aseg.deep.mgz file, prioritising reviewing the accuracy of the hippocampus segmentation. No datasets were removed for poor segmentation of the hippocampus. As FastSurfer outputs conform to Freesurfer’s recon-all outputs (58), the next Freesurfer’s hippocampal subfield segmentation analysis was conducted, with the addition of within-subject aligned T2-weighted scans to leverage multispectral segmentation improvements (Version 8.0.0; (59)). Following visual inspection of hippocampal subfields per participant, no datasets were excluded due to poor or unacceptable segmentations. Volume of hemisphere specific whole hippocampal structures, 19 hippocampal subfields segmentations and a single estimated total intracranial volume (eTIV) measure were extracted using in-house scripts. For the hippocampal subfield analyses, the cornu ammonis (CA) aggregation scheme was applied, resulting in 11 subfields per hemisphere: parasubiculum, presubiculum (combining presubiculum-head and presubiculum-body), subiculum (combining subiculum-head and subiculum-body), CA1 (combining CA1-head and CA1-body), CA3 (combining CA3-head and CA3-body), CA4 (combining CA4-head, CA4-body, GC-ML-DG-head, and GC-ML-DG-body), molecular layer (combining molecular_layer_HP-head and molecular_layer_HP-body), hippocampal-amygdala transition area (HATA), fimbria, hippocampal tail, and hippocampal fissure. All raw segmentation volumes underwent intracranial volume correction (ICV) using a residual approach based on individuals’ eTIV. All subsequent analyses utilising anatomical volumes reflect group-level residual corrected volumes.

#### 2.2.3 Exercise

Time engaged in exercise during a typical week was measured using the Godin Leisure-Time Exercise Questionnaire (60). This validated tool captures weekly frequency of strenuous, moderate, and light exercise activities lasting more than 15 minutes. Total activity scores were computed as: (9 × Strenuous) + (5 × Moderate) + (3 × Light), with weights representing approximate metabolic equivalents (METs) for each exercise intensity. Participants scores recorded as a continuous variable (non-categorised).

### 2.3 Statistical Analysis

#### 2.3.1 Data preparation

All statistical analyses were conducted using MATLAB R2024a. To address the positively skewed distribution typical of circulating biomarkers, irisin values were winsorized at ±3 standard deviations from the mean, following the established methodology for hormone biomarkers (61–63). This approach resulted in n=3 (4.1%) of values being capped at the upper threshold of 18.15 ng/mL (mean + 3×SD), with no adjustments required at the lower boundary. Irisin values were then natural log-transformed, with all subsequent analyses involving irisin using these Winsorized log-transformed values.

#### 2.3.2 Statistical Analyses

All analyses used fully standardised variables (z-scored) to facilitate effect size comparisons, with standardised beta coefficients (β) and 95% confidence intervals reported.

##### Hypothesis 1: Exercise levels are positively associated with circulating irisin

Hierarchical linear regression examined whether self-reported exercise (Godin Leisure-Time Exercise scores) predicted circulating log-transformed irisin levels. Age, sex, and years of education were entered as covariates in Step 1, with exercise added in Step 2 to assess its unique contribution (R^2^ change).

##### Hypothesis 2: Exercise levels are positively associated with hippocampal volumes

Hierarchical linear regression examined whether self-reported exercise predicted left and right hippocampal volumes separately. Age, sex, and years of education were entered as covariates in Step 1, with exercise added in Step 2.

##### Hypothesis 3: Irisin is positively associated with hippocampal volumes

Hierarchical linear regression examined associations between log-transformed irisin and hippocampal volumes at two anatomical levels. For all models, age, sex, and years of education were entered as covariates in Step 1, with log-transformed irisin added in Step 2.

We first tested associations with whole left and right hippocampal volumes (Level 1: Whole Structure; 2 tests). Next, we examined hippocampal subfields (Level 2: Hippocampal Subfields; 22 tests) by analysing eleven subfields bilaterally: parasubiculum, presubiculum, subiculum, CA1, CA3, CA4, molecular layer, HATA, fimbria, hippocampal tail, and hippocampal fissure. Benjamini-Hochberg False Discovery Rate (FDR) correction was applied separately at each level to control Type I error.

To test regional specificity, we conducted a control analysis using the primary visual cortex (bilateral pericalcarine regions averaged) with the same hierarchical regression approach. Effect sizes were quantified using Cohen’s f^2^ (R^2^change / (1 – R^2^full)) with interpretive thresholds of f^2^ ≥ 0.02 (small), ≥ 0.15 (medium), and ≥ 0.35 (large) (64, 65).

##### Post-Hoc tests of lateralisation

To examine potential hemispheric differences in irisin-hippocampus associations, we conducted post-hoc analyses on bilateral hippocampal subfield pairs. While our main hierarchical analysis identified nine individual hippocampal subfields with significant irisin associations, these represented six unique anatomical structures measured bilaterally (CA1, CA3, CA4, molecular layer, HATA, and fimbria). All variables were first standardised (z-scored). Age, sex, and education were then regressed from both the log-transformed irisin values and from each hippocampal subfield volume, with the resulting residuals used for correlation analyses. This approach ensured that any observed hemispheric differences are not driven by demographic factors and replicates our Hierarchical Regression analyses.

Steiger’s Z test for dependent correlations with overlapping variables (66) was implemented using the *cocor* package in R (67) to test whether the correlation between irisin and the right hemisphere subfield differed significantly from the correlation between irisin and the corresponding left hemisphere subfield. This test appropriately accounts for the dependency between measurements, including the within-subject correlation between left and right hippocampal volumes. FDR correction was applied across the six tested bilateral pairs to control for multiple comparisons, with significance set at FDR-p < 0.05.

##### Power Analysis

Post-hoc power analyses were conducted using G*Power 3.1.9.7 (68) for F-tests (Linear multiple regression: Fixed model, R^2^ increase) with one tested predictor and four total predictors. Power was calculated for observed effect sizes (Cohen’s f^2^) at α=0.05 and α=0.002 (FDR-corrected for 22 comparisons).

##### Hypothesis 4: Irisin mediates the relationship between exercise and hippocampal volumes

Following established mediation methodology (69, 70), for hippocampal subfields showing significant irisin-hippocampus associations in Hypothesis 3, we tested whether irisin mediates the exercise-hippocampus relationship using the Mediation Toolbox [https://github.com/canlab/MediationToolbox] in MATLAB, as per previous research (71–73). All variables were standardised (z-scored) to obtain standardised beta coefficients, with irisin log-transformed prior to standardisation to match the regression analyses. The model examined: (a) exercise→irisin effect, (b) irisin→hippocampus subfield of interest effect controlling for exercise, (c’) direct exercise→hippocampus subfield of interest effect controlling for irisin, (c) total exercise→hippocampus subfield of interest effect, and (ab) indirect effect of exercise through irisin. Analyses employed bias-corrected bootstrap procedures (5,000 iterations) to compute 95% confidence intervals and p-values. Age, sex, and education were standardised and included in each model as covariates. FDR correction was applied to indirect effects across all tested regions. Significance was set at p<0.05 (FDR-corrected) for all analyses.

## 3 RESULTS

### 3.1 Exercise-Irisin-Hippocampal Volume Regressions

#### 3.1.1 Exercise positively associated with irisin

Hierarchical regression analysis examined the relationship between exercise and irisin levels. Step 1 included demographic covariates (age, sex, education) and was not significant (F(3,70) = 0.153, p = 0.928, R^2^ = 0.007, Adjusted R^2^ = -0.036). Step 2 added exercise as a predictor, significantly improving model fit (R^2^ = 0.124, Adjusted R^2^ = 0.074). Exercise was a significant predictor (β = 0.365, 95% CI [0.130, 0.599], p = 0.003, Figure 1A), explaining an additional 11.8% of variance beyond covariates (ΔR^2^ = 0.118, Cohen’s f^2^ = 0.134).

**Figure 1:**
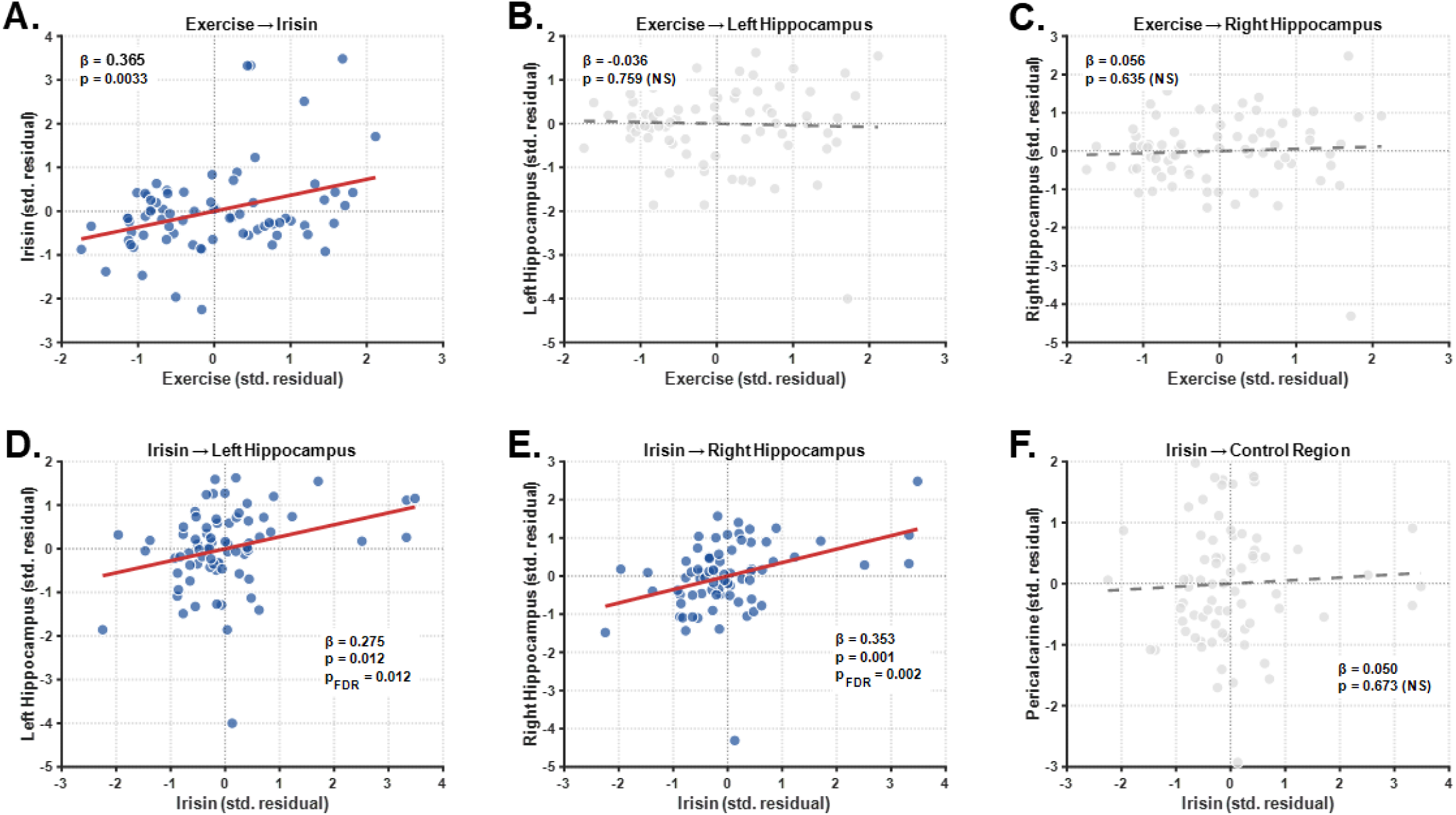
Exercise-irisin-hippocampus regression analysis. Scatter plots show associations between **(A)** self-reported exercise and circulating irisin levels, **(B)** exercise and left-hemisphere hippocampal volume, **(C)** exercise and right-hemisphere hippocampal volume, **(D)** irisin and left-hemisphere hippocampal volume, **(E)** irisin and right-hemisphere hippocampal volume, and **(F)** irisin and pericalcarine cortex volume (control region). Each point represents one participant (N = 74). Solid red lines indicate significant positive associations. Dashed gray lines indicate non-significant relationships. Beta coefficients and p-values are displayed on each panel. NS = non-significant. All variables shown are standardised residuals after adjusting for age, sex, and education. All axes represent standard deviation units centered at zero.

#### 3.1.2 Exercise did not associate with whole structure hippocampal volumes

For the left hippocampus, Step 1 with demographic covariates was significant (F(3,70) = 3.952, p = 0.012, R^2^ = 0.145, Adjusted R^2^ = 0.108). Step 2 added exercise, which did not improve model fit (R^2^ = 0.146, Adjusted R^2^ = 0.097). Exercise was not a significant predictor (β = -0.036, 95% CI [-0.268, 0.195], p = 0.759, ΔR^2^ = 0.001, Cohen’s f^2^ = 0.001, Figure 1B). For the right hippocampus, Step 1 was significant (F(3,70) = 4.096, p = 0.010, R^2^ = 0.149, Adjusted R^2^ = 0.113). Step 2 with exercise showed no improvement (R^2^ = 0.152, Adjusted R^2^ = 0.103), with exercise not significantly predicting volume (β = 0.056, 95% CI [-0.175, 0.287], p = 0.635, ΔR^2^ = 0.003, Cohen’s f^2^ = 0.003, Figure 1C).

#### 3.1.3 Irisin positively associated with whole hippocampal structural volumes

Hierarchical regression analysis examined the relationship between irisin and hippocampal volumes bilaterally. For the left hippocampus, Step 1 with demographic covariates was significant (F(3,70) = 3.952, p = 0.012, R^2^ = 0.145, Adjusted R^2^ = 0.108). Step 2 added irisin, significantly improving model fit (R^2^ = 0.220, Adjusted R^2^ = 0.175). Irisin was a significant predictor (β = 0.275, 95% CI [0.066, 0.484], p = 0.012, FDR-p = 0.012, Figure 1D), explaining an additional 7.5% of variance (ΔR^2^ = 0.075, Cohen’s f^2^ = 0.096).

For the right hippocampus, Step 1 was significant (F(3,70) = 4.096, p = 0.010, R^2^ = 0.149, Adjusted R^2^ = 0.113). Step 2 with irisin showed significant improvement (R^2^ = 0.273, Adjusted R^2^ = 0.231). Irisin significantly predicted volume (β = 0.353, 95% CI [0.151, 0.555], p = 0.001, FDR-p = 0.002, Figure 1E), explaining an additional 12.4% of variance (ΔR^2^ = 0.124, Cohen’s f^2^ = 0.170).

#### 3.1.4 Irisin did not associate with a control brain region volume

Hierarchical regression analysis examined the relationship between irisin and pericalcarine cortex volume as a control region. Step 1 with demographic covariates was not significant (F(3,70) = 0.957, p = 0.418, R^2^ = 0.039, Adjusted R^2^ = -0.002). Step 2 added irisin, showing no improvement in model fit (R^2^ = 0.042, Adjusted R^2^ = -0.014). Irisin did not predict pericalcarine volume (β = 0.050, 95% CI [-0.182, 0.282], p = 0.673, Figure 1F), with negligible variance explained (ΔR^2^ = 0.002, Cohen’s f^2^ = 0.003).

### 3.2 Hippocampal subfields and irisin relationships

#### 3.2.1 Hippocampal Subfield Analysis

Hierarchical regression analyses performed across all hippocampal subfields revealed heterogeneous associations with irisin. After FDR correction for multiple comparisons, nine out of 22 hippocampal subfields models were significant, all having positive associations with irisin levels (Supplementary Table 1, Figure 2).

**Figure 2.**
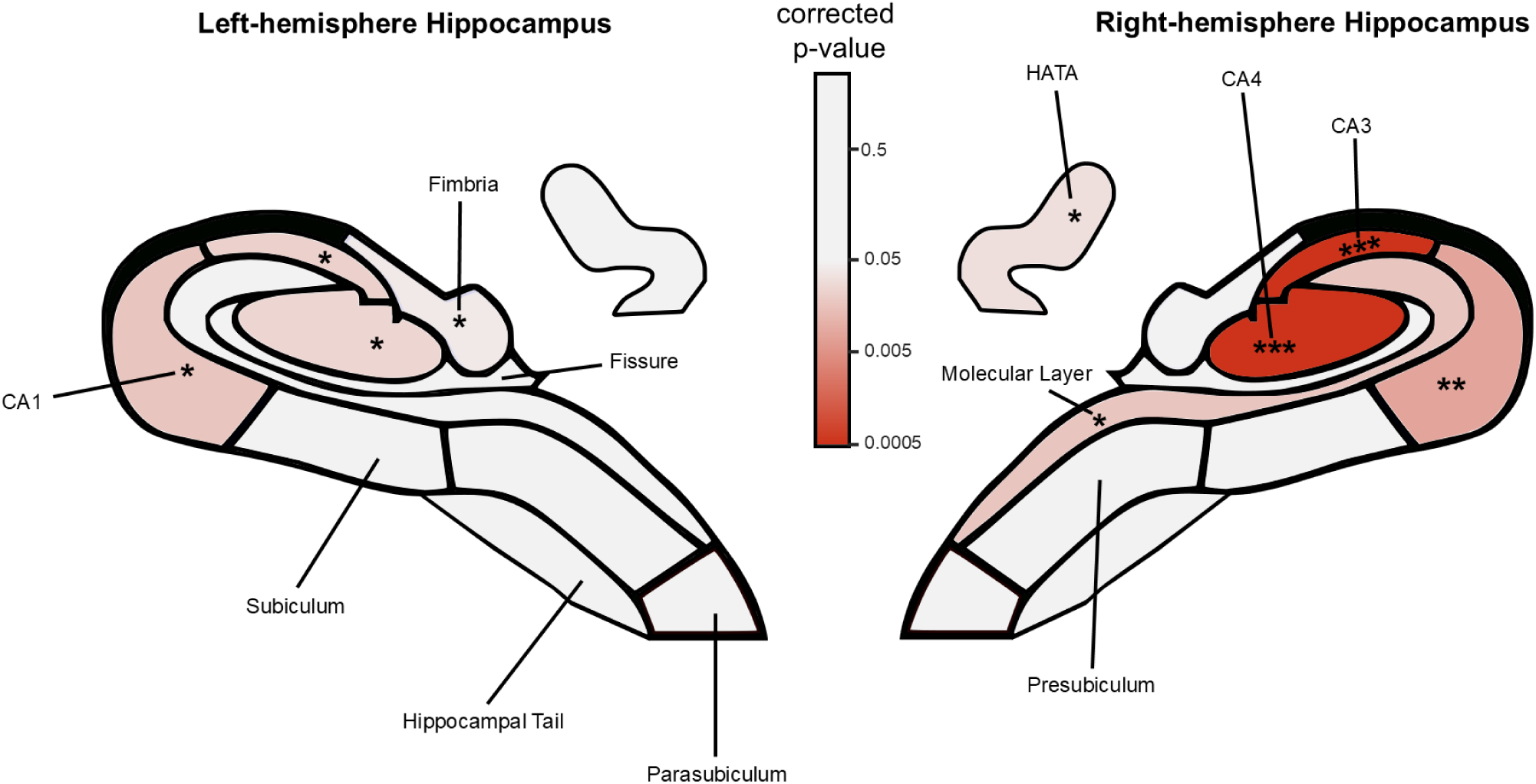
Hippocampal subfield map showing statistical significance of irisin associations. Colour-coded representation of FDR-corrected p-values for the association between circulating irisin levels and hippocampal subfield volumes from hierarchical regression analysis (Step 2 beta coefficient) after controlling for age, sex, and education (Step 1). The colour gradient represents statistical significance, with warmer colours (red) indicating stronger associations (lower p-values) and cooler colours (pink to white) indicating weaker or non-significant associations (p ≥ 0.05). All significant associations were positive, indicating higher irisin levels were associated with larger subfield volumes. Nine of 22 subfield models showed significant positive associations. Asterisks (*) indicate subfields with Benjamini-Hochberg FDR-p < 0.05; double asterisks (**) indicate FDR-p < 0.01; triple asterisks (***) indicate FDR-p < 0.001. Visual depiction of hippocampal subfield outlines were modified from (74). CA, cornu ammonis; HATA, hippocampus-amygdala transition area.

#### 3.2.2 Hemispheric Asymmetry in Irisin-Hippocampus Associations

Post-hoc analyses examined hemispheric differences in the six subfields showing significant associations with irisin. After regressing out age, sex, and education, Steiger’s Z tests for dependent correlations revealed significant hemispheric asymmetry in CA3, with stronger irisin associations in the right hemisphere (r RH = 0.547, r LH = 0.323, difference = 0.224, Z = 2.85, p = 0.004, FDR-p = 0.026). CA4 showed a trend toward right hemispheric asymmetry (r RH = 0.490, r LH = 0.312, difference = 0.178, Z = 2.36, p = 0.018, FDR-p = 0.055). The remaining subfields showed no significant hemispheric differences after FDR correction (all FDR-p > 0.14).

#### 3.2.3 Post-hoc Power Analysis

Power analysis revealed our sample (N=74) achieved 94% power for detecting medium-to-large effects (f^2^=0.17). For the nine significant hippocampal subfield associations, we achieved power ranging from 70.9% (fimbria left, f^2^=0.088) to >99.9% (CA3 right, f^2^=0.426). After FDR correction (α=0.002), power was 64.6% for medium effects and 98.8% for large effects.

### 3.3 Mediation Analysis

Following methodological recommendations for testing indirect pathways (69, 70, 75), we proceeded with mediation analysis to test whether circulating irisin provides an indirect pathway linking exercise to hippocampal volumes in our sample.

The exercise-irisin association (Figure 3, path a) was consistent across all models (Table 2, β = 0.365, p < 0.001). The irisin-hippocampus associations (Figure 3, path b) were significant for all regions tested (Table 2, FDR-p < 0.05). Critically, all indirect effects (Figure 3, path ab) were significant after FDR correction (Table 2, all FDR-p ≤ 0.006), with the strongest mediation observed in right CA3 (Table 2, β-ab = 0.201, 95% CI [0.153, 0.274], FDR-p < 0.001) and right CA4 (Table 2, β-ab = 0.186, 95% CI [0.145, 0.255], FDR-p < 0.001). All nine regions demonstrated significant indirect effects through irisin (path ab), despite non-significant total effects (path c) and direct effects (path c’, all ps ≥ 0.305), representing indirect-only mediation where exercise relates to hippocampal volumes exclusively through the irisin pathway. The consistent positive direction of all paths (exercise→irisin and irisin→hippocampus) indicates consistent rather than inconsistent mediation (69), suggesting a unified mechanism rather than opposing processes.

**Figure 3.**
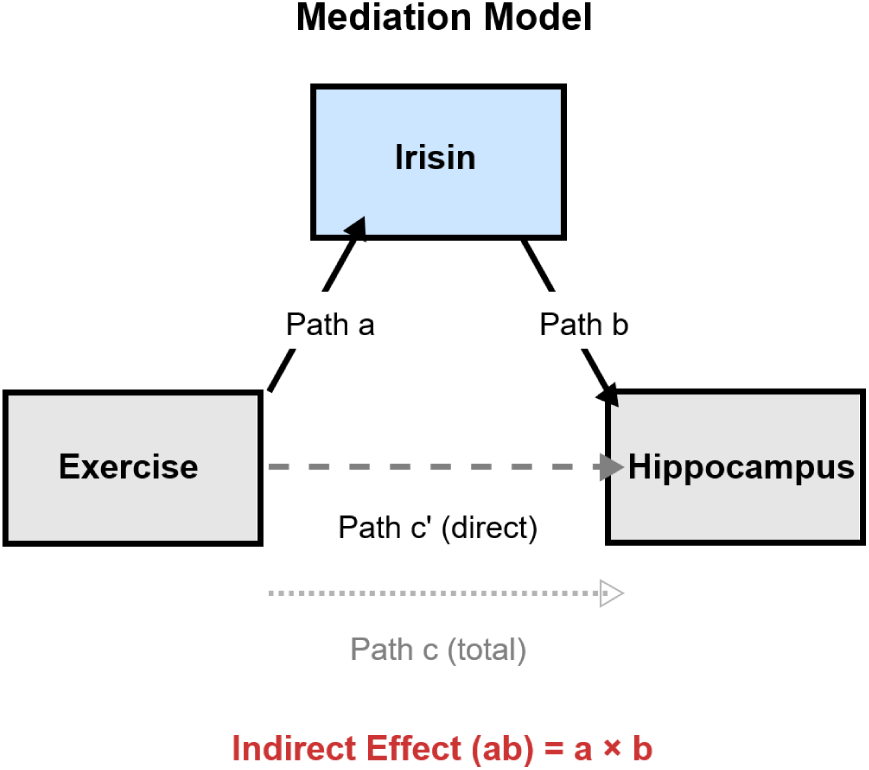
Mediation Analysis Structure. In our mediation analysis, *path a* represents the effect of exercise on irisin, *path b* represents the effect of irisin on hippocampal volumes (controlling for exercise), *path c* represents the total effect of exercise on hippocampal volumes, *path c’* represents the direct effect (controlling for irisin), and *path ab* represents the indirect effect of exercise on hippocampal volumes through irisin.

**Table 1.** Description of Sample.

| Measure | Number | Mean (standard deviation) | Range (min-max) |
| --- | --- | --- | --- |
| Age | 74 | 65.47 (8.56) | 50.15 – 84.04 |
| Sex (F/M) | 74 | (61/13) |  |
| Education<sup>a</sup> | 74 | 14.74 years (2.83) | 9 - 19 |
| Self-Reported Exercise METS<sup>b</sup> | 74 | 42.5 (24.08) | 0-101 |
| Irisin | 74 | 5.93 (2.97) | 2.45-18.15 |
| Total Hippocampal Volume<sup>c</sup> | 74 | 6139.07 (825.52) | 4188.37-7920.72 |
| Total Pericalcarine Volume<sup>c</sup> | 74 | 1826.41 (395.00) | 776.00-2571.00 |

<sup>a</sup> Self-reported in years
<sup>b</sup> Metabolic Equivalent of Task units from Godin Leisure-Time Exercise Questionnaire;
<sup>c</sup> Values are in raw units (mm<sup>3</sup>)

**Table 2.** Mediation Results.

| Region | $\beta$ path<br>a | p-value<br>path a | $\beta$ path<br>b | p-value<br>path b | $\beta$ path<br>c' | p-value<br>path c' | $\beta$ path ab<br>[95% CI] | p-value<br>path ab | FDR-p<br>path ab |
| --- | --- | --- | --- | --- | --- | --- | --- | --- | --- |
| CA3 Right | **0.365** | **<0.001** | **0.551** | **<0.001** | -0.065 | 0.526 | **0.201**<br>[0.153, 0.274] | **<0.001** | **<0.001** |
| CA4 Right | **0.365** | **<0.001** | **0.509** | **<0.001** | -0.119 | 0.521 | **0.186**<br>[0.145, 0.255] | **<0.001** | **<0.001** |
| CA1 Right | **0.365** | **<0.001** | **0.396** | **<0.001** | -0.071 | 0.698 | **0.144**<br>[0.106, 0.200] | **<0.001** | **<0.001** |
| CA1 Left | **0.365** | **<0.001** | **0.360** | **0.001** | -0.076 | 0.682 | **0.131**<br>[0.099, 0.171] | **<0.001** | **<0.001** |
| Molecular Right | **0.365** | **<0.001** | **0.340** | **0.006** | -0.095 | 0.652 | **0.124**<br>[0.089, 0.177] | **<0.001** | **<0.001** |
| CA3 Left | **0.365** | **<0.001** | **0.338** | **0.004** | -0.064 | 0.609 | **0.123**<br>[0.093, 0.174] | **0.002** | **0.002** |
| CA4 Left | **0.365** | **<0.001** | **0.332** | **0.005** | -0.116 | 0.453 | **0.121**<br>[0.090, 0.170] | **0.002** | **0.002** |
| Fimbria Left | **0.365** | **<0.001** | **0.288** | **0.006** | -0.120 | 0.305 | **0.105**<br>[0.074, 0.156] | **<0.001** | **<0.001** |
| HATA Right | **0.365** | **<0.001** | **0.280** | **0.022** | 0.025 | 0.746 | **0.102**<br>[0.072, 0.153] | **0.006** | **0.006** |

## 4 DISCUSSION

### 4.1 Main Findings

We provide the first human evidence that circulating irisin mediates the relationship between exercise and hippocampal volume. In our cross-sectional sample of 74 healthy older adults, exercise was positively associated with circulating irisin levels (β=0.365, p=0.003), and irisin levels were positively associated with bilateral whole-structure hippocampal volumes (left: 7.5% unique variance, FDR-p=0.012; right: 12.4% unique variance, FDR-p=0.002). This relationship demonstrated anatomical specificity, as irisin showed no association with our control brain region (pericalcarine cortex, p=0.673). Further analyses revealed irisin’s selective relationships within hippocampal subfields. The strongest relationships were in CA3 (right: 27.9% variance explained; left: 10.0%), CA4 (right: 22.0%; left: 8.6%), and CA1 (right: 13.8%; left: 11.2%), alongside weaker relationships in right molecular layer, right HATA, and left fimbria. Post-hoc analyses revealed significant hemispheric asymmetry in irisin’s relationships favouring the right hemisphere for both CA3 (Z=2.85, FDR-p=0.026) and trending for CA4 (Z=2.36, FDR-p=0.055). Notably, our mediation analyses revealed significant indirect effects of exercise on hippocampal volumes through irisin across all nine hippocampal subfields identified from our regression analysis (indirect effects β∼0.10-0.20, all FDR-corrected p≤0.006), with no significant direct effects of exercise on hippocampal volumes when controlling for irisin (all path c’ p≥0.305). Overall, these results position irisin as a critical molecular mediator between exercise and hippocampal volumes in older adults.

### 4.2 Irisin as a Molecular Mechanism for Exercise-Hippocampus Effects

Exercise is increasingly recognised as a powerful non-pharmacological intervention for mitigating age and disease-related brain decline (16). The hippocampus is particularly responsive, with studies demonstrating that exercise can both preserve and increase older adults’ hippocampal volumes (20–22, 76). The myokine irisin has recently emerged as a possible mechanism underlying these benefits (25, 28, 30). Indeed, animal models have demonstrated that irisin is critical for exercise-induced improvements in hippocampal plasticity and neurogenesis (38, 39, 42), acting through multiple converging neuroprotective mechanisms, reviewed in significant detail elsewhere (28, 30).

Crucially, while previous human studies have demonstrated that exercise elevates circulating irisin (43–47) and that irisin positively correlates with cognitive function (50, 52, 77), direct evidence linking circulating irisin to hippocampal structure was absent. Our analysis now provides direct human evidence for this pathway, demonstrating significant indirect effects of exercise on hippocampal volumes through irisin in a pattern consistent with the molecular mechanisms identified in animal models.

Importantly, these findings may help explain the specific hippocampal changes observed in previous exercise intervention studies. For example, Erickson et al. (2011) found that one year of walking intervention increased hippocampal volume of older adults, with effects concentrated in the subiculum, CA1, and dentate gyrus. Our results offer a mechanistic explanation for some of this regional specificity, showing that exercise-related circulating irisin is associated with the volumes of CA1 and CA4 (which included the dentate gyrus). Interestingly, we found no irisin-subiculum association, suggesting irisin-independent pathways also contribute to exercise’s hippocampal benefits. Importantly, our finding of non-significant total effects does not contradict the substantial literature demonstrating direct exercise-hippocampus relationships (17, 18, 20–22). Rather, the detection of significant indirect effects through irisin, despite non-significant total effects in our sample, identifies irisin as an important, but not exclusive mechanism through which exercise influences hippocampal volume.

### 4.3 Implications for Brain Ageing and Preventing Neurodegenerative Disease

We found no relationship between irisin and the subiculum, the subfield most susceptible to decline in normal ageing (3, 78, 79). In contrast, irisin showed strong bilateral associations with CA1, a hippocampal subfield that experiences the earliest and most severe atrophy in AD while remaining relatively preserved in healthy ageing (78, 80–84). This indicates irisin may preferentially support regions vulnerable to pathological rather than normal age-related changes.

Beyond CA1, we observed strong bilateral associations between irisin and both CA3 and CA4, subfields critical for memory function (85, 86) that show variable vulnerability patterns in ageing and disease. While some studies report preservation in both AD and healthy ageing (83, 84), others document significant atrophy in AD (87), with considerable heterogeneity noted across the literature (3). This variability in CA3 and CA4 vulnerability suggests that pathological susceptibility alone cannot explain irisin’s hippocampal associations. Notably, our CA4 segmentation includes the dentate gyrus, where adult neurogenesis occurs (88, 89). Given that exercise-induced irisin promotes neurogenesis in animal models (40, 41), with its effects observed in the dentate gyrus of rodents (40), our irisin-CA4 associations may therefore reflect irisin supporting neurogenic processes in this subfield.

The additional, albeit weaker, associations with right molecular layer, right HATA, and left fimbria show irisin’s relationships extend beyond the core CA subfields. These associations may reflect a ’neighbourhood effect’ given these structures are anatomically adjacent to or directly connected with the strongly associated CA subfields (see Figure 2), unlike more spatially distant regions, such as the parasubiculum or hippocampal tail, which showed no associations with irisin.

Interestingly, our results contrast with Kim et al. (2022), who found no irisin-hippocampus relationship in healthy controls (n=23) using total hemisphere volumes. Our study revealed strong positive associations, likely due to our increased statistical power from our larger healthy sample (n=74) alongside subfield-specific analyses. Kim et al. (2022) also observed a reversal in AD patients—higher plasma irisin correlated with smaller hippocampal volumes, interpreted as "irisin resistance” (51). This may reflect a critical transition where irisin supports hippocampus volume in healthy ageing, but not in AD, where elevated levels may indicate unsuccessful compensation.

### 4.4 Hemispheric Differences

Interestingly, direct hemispheric comparison revealed that right CA3 and right CA4 showed stronger irisin associations than their left-hemisphere counterparts. This right-hemisphere predominance aligns with well-established patterns of hippocampal asymmetry in healthy ageing, where previous studies employing MRI-structural analysis have demonstrated consistent rightward volumetric asymmetry (right > left) that strengthens with age (12, 90). This asymmetry undergoes characteristic changes during pathological ageing.

Rightward lateralisation of hippocampal volume peaks in MCI before diminishing in AD as accelerated bilateral atrophy reduces the hemispheric imbalance (12, 91). While CA1 represents the earliest site of AD pathology (3), it along with CA3 and CA4 nonetheless maintain rightward lateralisation across healthy controls, MCI, and AD populations (92). Our finding that irisin preferentially associates with right CA3 and CA4, hemispheres that normally maintain greater volume, suggests irisin may support these lateralisation patterns across healthy and pathological ageing changes.

### 4.5 Limitations and Strengths

Several limitations should be considered when interpreting our findings. First, our cross-sectional study design cannot establish causality, though we used mediation analysis to explore potential mechanisms. The use of mediation on cross-sectional data is considered appropriate when the analysis is guided by strong theoretical foundations and serves as a more powerful test of a mechanism than an analysis of the total effect alone (69, 75, 93). Further, while our cross-sectional design cannot establish temporal precedence, the causal sequence is strongly supported by prior studies showing exercise precedes irisin elevation (43, 45, 47), and animal studies demonstrating irisin subsequently affects hippocampal health (38–40). Second, there is ongoing methodological debate regarding the cross-reactivity and specificity of commercial irisin immunoassays ((94), for review, see (95)), although our use of a single assay lot across all samples ensures that the relative differences driving our mediation effects remain valid regardless of absolute measurement accuracy. Third, although our use of the Godin questionnaire to quantify exercise introduces potential self-report biases, it has well-established validity with objective fitness measures (96). Finally, our focus on irisin excludes other potentially important exercise-induced factors that might jointly contribute to hippocampal volume. Other mechanisms, including BDNF upregulation, vascular changes, and additional myokines, likely contribute to the well-established exercise-hippocampus relationship. Rather, our results support irisin as a critical molecular pathway through which exercise might modify hippocampal volumes in older adults. Future longitudinal studies with objective activity monitoring and broader myokine panels will be crucial to fully elucidate these pathways.

Despite these limitations, our study has several notable strengths. The multimodal integration of behavioural, biochemical, and neuroimaging data allowed for the first demonstration in humans that circulating irisin mediates the relationship between exercise and hippocampal volumes. Our analytical approach strengthened mechanistic inferences by controlling for demographic factors that could confound the relationship between exercise, irisin, and brain structure (e.g., (97)), and presents strong anatomically specific relationships even after correction for multiple comparisons and demographic controls, highlighting their reliability. The specificity of our findings to hippocampal subfields, alongside null findings in control region analyses, further supports the validity of these associations.

### 4.6 Conclusions

This study provides the first human evidence that circulating irisin mediates the relationship between exercise and hippocampal volume in healthy older adults. The significant indirect pathway through irisin, demonstrated despite non-significant total effects, identifies a critical molecular pathway linking physical activity to brain structure. The anatomical specificity of these associations, strongest in cornu ammonis subfields (CA1, CA3, CA4) with right-hemisphere predominance, suggests irisin may preferentially support regions critical for memory, neurogenesis, and AD vulnerability. These findings have immediate translational implications where irisin-based interventions could amplify exercise benefits or provide alternative neuroprotection for mobility-limited older adults. Future longitudinal studies should examine whether maintaining optimal irisin levels through exercise or pharmacological means can prevent the transition from healthy to pathological brain ageing.

## Data Availability

The datasets presented within this study are available from the corresponding author on reasonable request.

## Acknowledgements

This study presents independent research supported by the Wilson Foundation.

## Conflicts of Interest

The authors declare no conflicts of interest.

## Notes

### Competing Interest Statement

The authors have declared no competing interest.

### Author Declarations

University of the Sunshine Coast's Human Research Ethics Committee (HREC) gave ethical approval for this work.

