## Supplementary Table 1 for "The myokine irisin represents an indirect pathway linking exercise to hippocampal subfields relevant to Alzheimer’s disease and neurogenesis"

**Supplementary Results**

**Supplementary Table 1. Irisin associations with hippocampal subfields**

| **Region** | **Beta** | **SE** | **CI 95** | **Raw p** | **FDR p** | **R^2^ change** | **Cohens f^2^** |
| --- | --- | --- | --- | --- | --- | --- | --- |
| CA1 Left | 0.336 | 0.111 | [0.117, 0.554] | 0.004 | **0.017** | 0.112 | 0.131 |
| CA1 Right | 0.373 | 0.109 | [0.159, 0.586] | 0.001 | **0.008** | 0.138 | 0.169 |
| CA3 Left | 0.317 | 0.112 | [0.098, 0.537] | 0.006 | **0.022** | 0.100 | 0.117 |
| CA3 Right | 0.530 | 0.098 | [0.338, 0.721] | <0.001 | **<0.001** | 0.279 | 0.426 |
| CA4 Left | 0.294 | 0.108 | [0.083, 0.506] | 0.008 | **0.025** | 0.086 | 0.108 |
| CA4 Right | 0.471 | 0.101 | [0.273, 0.668] | <0.001 | **<0.001** | 0.220 | 0.316 |
| HATA Left | 0.164 | 0.112 | [-0.055, 0.383] | 0.146 | 0.196 | 0.027 | 0.031 |
| HATA Right | 0.288 | 0.111 | [0.071, 0.504] | 0.011 | **0.031** | 0.082 | 0.098 |
| Hippocampal tail Left | 0.091 | 0.113 | [-0.129, 0.312] | 0.420 | 0.486 | 0.008 | 0.010 |
| Hippocampal tail Right | 0.172 | 0.112 | [-0.046, 0.391] | 0.127 | 0.196 | 0.029 | 0.035 |
| fimbria Left | 0.249 | 0.101 | [0.051, 0.448] | 0.016 | **0.040** | 0.062 | 0.088 |
| fimbria Right | 0.165 | 0.114 | [-0.058, 0.388] | 0.151 | 0.196 | 0.027 | 0.031 |
| hippocampalfissure Left | 0.023 | 0.110 | [-0.192, 0.238] | 0.833 | 0.833 | 0.001 | 0.001 |
| hippocampalfissure Right | 0.131 | 0.113 | [-0.090, 0.353] | 0.248 | 0.303 | 0.017 | 0.020 |
| molecular Left | 0.230 | 0.109 | [0.016, 0.444] | 0.039 | 0.085 | 0.053 | 0.064 |
| molecular Right | 0.310 | 0.103 | [0.107, 0.512] | 0.004 | **0.017** | 0.095 | 0.130 |
| parasubiculum Left | 0.064 | 0.120 | [-0.172, 0.299] | 0.599 | 0.627 | 0.004 | 0.004 |
| parasubiculum Right | -0.064 | 0.115 | [-0.290, 0.162] | 0.582 | 0.627 | 0.004 | 0.004 |
| presubiculum Left | 0.200 | 0.109 | [-0.013, 0.413] | 0.070 | 0.118 | 0.040 | 0.049 |
| presubiculum Right | 0.194 | 0.098 | [0.002, 0.386] | 0.051 | 0.096 | 0.037 | 0.057 |
| subiculum Left | 0.165 | 0.114 | [-0.058, 0.388] | 0.152 | 0.196 | 0.027 | 0.030 |
| subiculum Right | 0.219 | 0.111 | [0.001, 0.436] | 0.052 | 0.096 | 0.048 | 0.056 |
